# Sex-Specific Skeletal Muscle Gene Expression Responses to Exercise Reveal Novel Direct Mediators of Insulin Sensitivity Change

**DOI:** 10.1101/2024.09.07.24313236

**Authors:** S Ma, MC Morris, MJ Hubal, LM Ross, KM Huffman, CG Vann, N Moore, ER Hauser, A Bareja, R Jiang, E Kummerfeld, MD Barberio, JA Houmard, WB Bennett, JL Johnson, JA Timmons, G Broderick, VB Kraus, CF Aliferis, WE Kraus

**Author notes:** Corresponding author: William E. Kraus, Duke Molecular Physiology Institute 300 N. Duke St., Durham, NC 27701. Equal contribution (S Ma, MC Morris).

## Abstract

**BACKGROUND:** Understanding the causal pathways, systems, and mechanisms through which exercise impacts human health is complex. This study explores molecular signaling related to whole-body insulin sensitivity (Si) by examining changes in skeletal muscle gene expression. The analysis considers differences by biological sex, exercise amount, and exercise intensity to identify potential molecular targets for developing pharmacologic agents that replicate the health benefits of exercise.

**METHODS:** The study involved 53 participants from the STRRIDE I and II trials who completed eight months of aerobic training. Skeletal muscle gene expression was measured using Affymetrix and Illumina technologies, while pre- and post-training Si was assessed via an intravenous glucose tolerance test. A novel gene discovery protocol, integrating three literature-derived and data-driven modeling strategies, was employed to identify causal pathways and direct causal factors based on differentially expressed transcripts associated with exercise intensity and amount.

**RESULTS:** In women, the transcription factor targets identified were primarily influenced by exercise amount and were generally inhibitory. In contrast, in men, these targets were driven by exercise intensity and were generally activating. Transcription factors such as ATF1, CEBPA, BACH2, and STAT1 were commonly activating in both sexes. Specific transcriptional targets related to exercise-induced Si improvements included TACR3 and TMC7 for intensity-driven effects, and GRIN3B and EIF3B for amount-driven effects. Two key signaling pathways mediating aerobic exercise-induced Si improvements were identified: one centered on estrogen signaling and the other on phorbol ester (PKC) signaling, both converging on the epidermal growth factor receptor (EGFR) and other relevant targets.

**CONCLUSIONS:** The signaling pathways mediating Si improvements from aerobic exercise differed by sex and were further distinguished by exercise intensity and amount. Transcriptional adaptations in skeletal muscle related to Si improvements appear to be causally linked to estrogen and PKC signaling, with EGFR and other identified targets emerging as potential skeletal muscle-specific drug targets to mimic the beneficial effects of exercise on Si.

## INTRODUCTION

Exercise training provides substantial health benefits; however, too few individuals adopt and maintain it as a lifelong health strategy ^1^. Consequently, there is significant interest in developing pharmacologic alternatives able to replicate the health effects of exercise ^2^. To facilitate such pharmacologic development, a deeper understanding is needed of the complex, pleiotropic, and sex-specific physiological effects of exercise, which occur across multiple organ systems. Identifying specific molecular mediators and causal pathways that connect specific exercise regimens to specific health outcomes is a crucial step in developing therapeutics that mimic the effects of exercise.

Given the substantial role of whole-body insulin action as a marker and mediator of cardiometabolic risk and dysfunction, skeletal muscle insulin sensitivity (insulin sensitivity index, Si) presents a potentially powerful pharmaceutical target. Extensive literature in both humans and animals indicates that exercise-induced improvements in whole-body insulin sensitivity are closely linked to molecular processes and adaptations in skeletal muscle ^3–6^. Based on this rationale, the purpose of this study is to causally model the effects of aerobic exercise training on the skeletal muscle transcriptome and its functional relationship to changes in Si ^7^.

This study leverages data from the STRRIDE (Studies of Targeted Risk Reduction Interventions through Defined Exercise) series, which examined the effects of varying amounts, intensities, and modes of exercise training on cardiometabolic disease risk factors ^8–11^. Conducted between 1998 and 2013, the three STRRIDE studies were designed to explore the temporal effects of eight months of exercise training and subsequent detraining on key clinical cardiometabolic and physiological outcomes. They also investigated the dose-response and mode-specific effects of exercise on these outcomes, with a focus on the molecular mechanisms in skeletal muscle mediating these effects. These studies have produced a robust repository of demographic, clinical, and molecular data from 920 enrollees and 580 completers across the three cohorts.

For this study, we utilized data from a subset of participants who completed aerobic exercise training in STRRIDE I and II. We hypothesized that transcription factor targets would be influenced by biological sex and specific parameters of the exercise training programs (e.g., amount and intensity). To identify regulatory and regulated elements leading to exercise-induced changes in Si, we employed two complementary approaches: integrative molecular physiology and advanced machine learning and causal discovery methods.

## RESULTS

### Transcription Factor Identification

We used PASTAA to predict transcription factors that regulate a set of genes based on annotated affinities calculated from biophysical interactions. Among the muscle genes that varied significantly by sex and/or exercise protocol parameters (intensity or amount), the PASTAA query identified 30 transcription factors predicted to be associated with the selected genes (p<0.05) (Supplemental Figure 1).

### Sex-Specificity of Exercise Amount and Intensity Transcription Factor Targets

To examine the sex-specific effects of exercise amount and intensity, we stratified the analyses by sex and controlled for sex in subsequent analyses. A total of 6,078 probes showed significant variation in expression across groups of men and women (uncorrected p<0.05). The cluster patterns resulting from multidimensional scaling of these probes indicated that exercise intensity, amount, and biological sex all significantly influenced the transcriptomic response (Figures 2, 3). The greatest divergence in co-expression profiles between men and women occurred where the effects of exercise amount and intensity were most pronounced.

**Figure 1.**
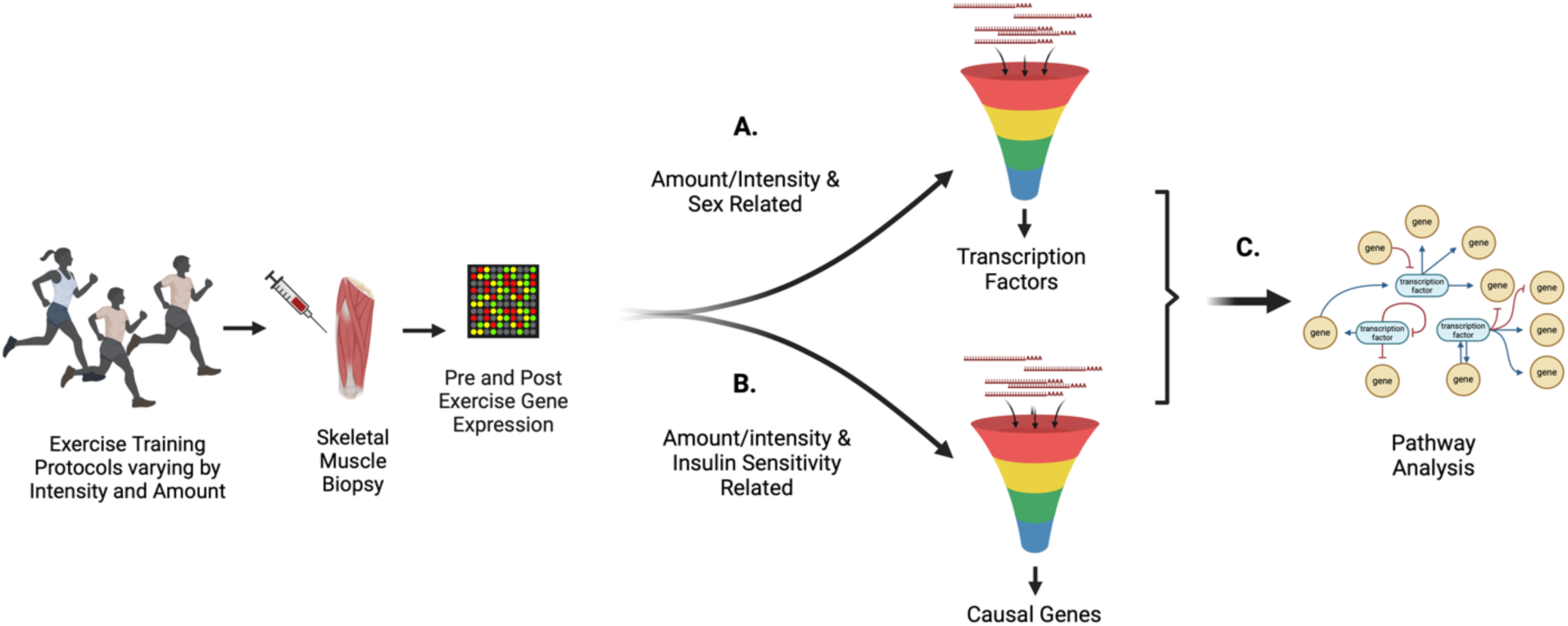
Conceptual Design. A candidate list of gene transcription targets was developed through three approaches: A. identification of a set of gene transcription factors modified by exercise depending on amount and intensity of exercise and biological sex constrained by prior knowledge; B. identification of a set of genes causally related to and constrained by their relation to change in insulin sensitivity as defined by the insulin sensitivity index; C. identification of the intersection of these two gene sets were used to identify gene expression networks and gene regulatory nodes causally connecting exercise of different amounts and intensities to changes in insulin sensitivity. Created in BioRender. Kraus, W. (2024) BioRender.com/t18t722.

**Figure 2.**
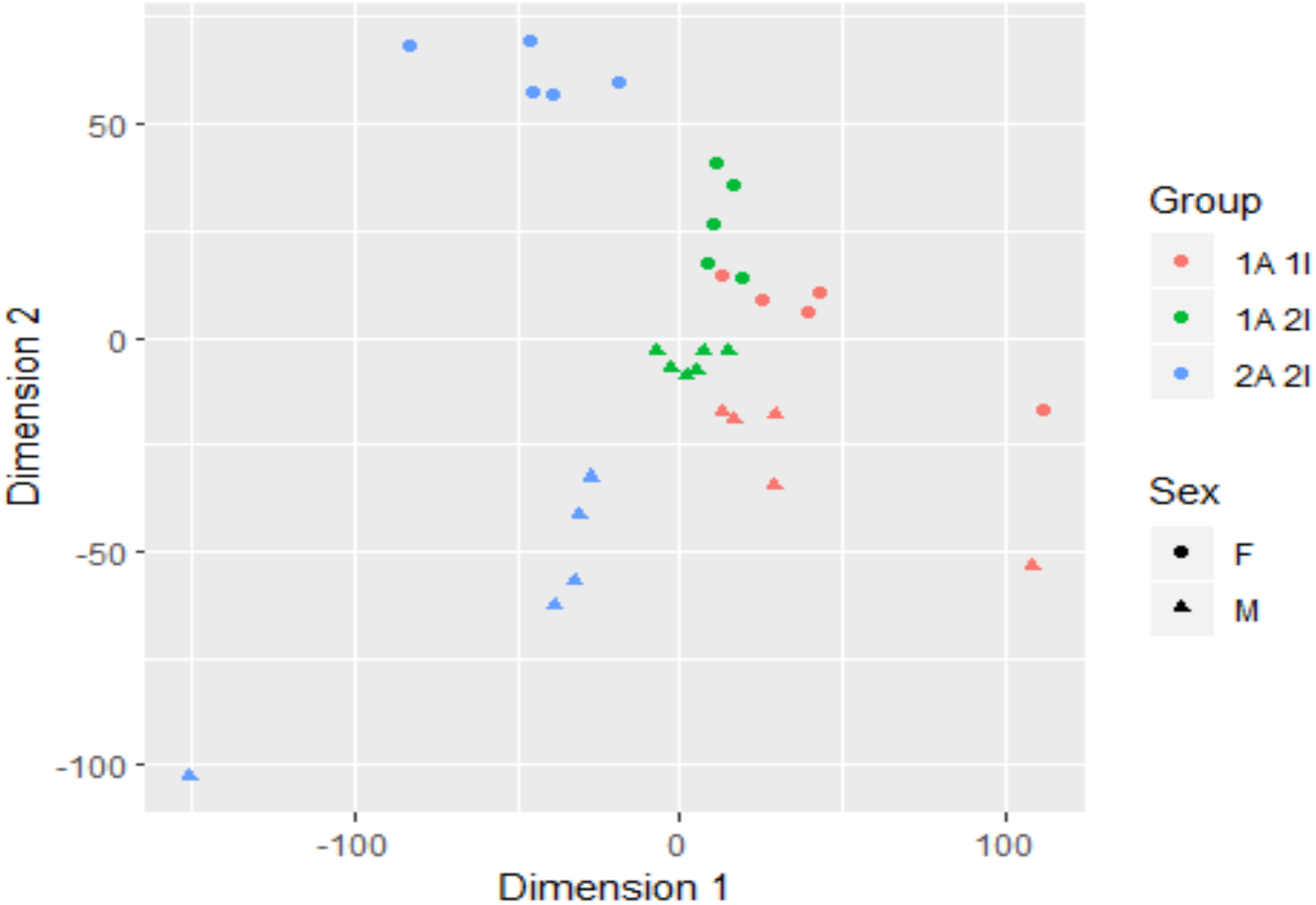
Multidimensional scaling (cluster analysis) of pre- to post-exercise change of transcriptional factors with significant variation according to participant sex (M, W) and/or exercise parameter (amount or intensity). The three colors – red, green and blue – are assigned to the different exercise groups, where “1A” and “2A” represent different amounts controlling for intensity, and “1I” and “2I” represent different intensities controlling for amount. The women are represented by filled circles and the men by filled triangles. Each point represents one participant. Overall transcriptomic responses in subjects correlated with both exercise protocol and biological sex. Dimension 1 separates by exercise groups characterized by intensity and amount; Dimension 2 separates by biological sex.

**Figure 3.**
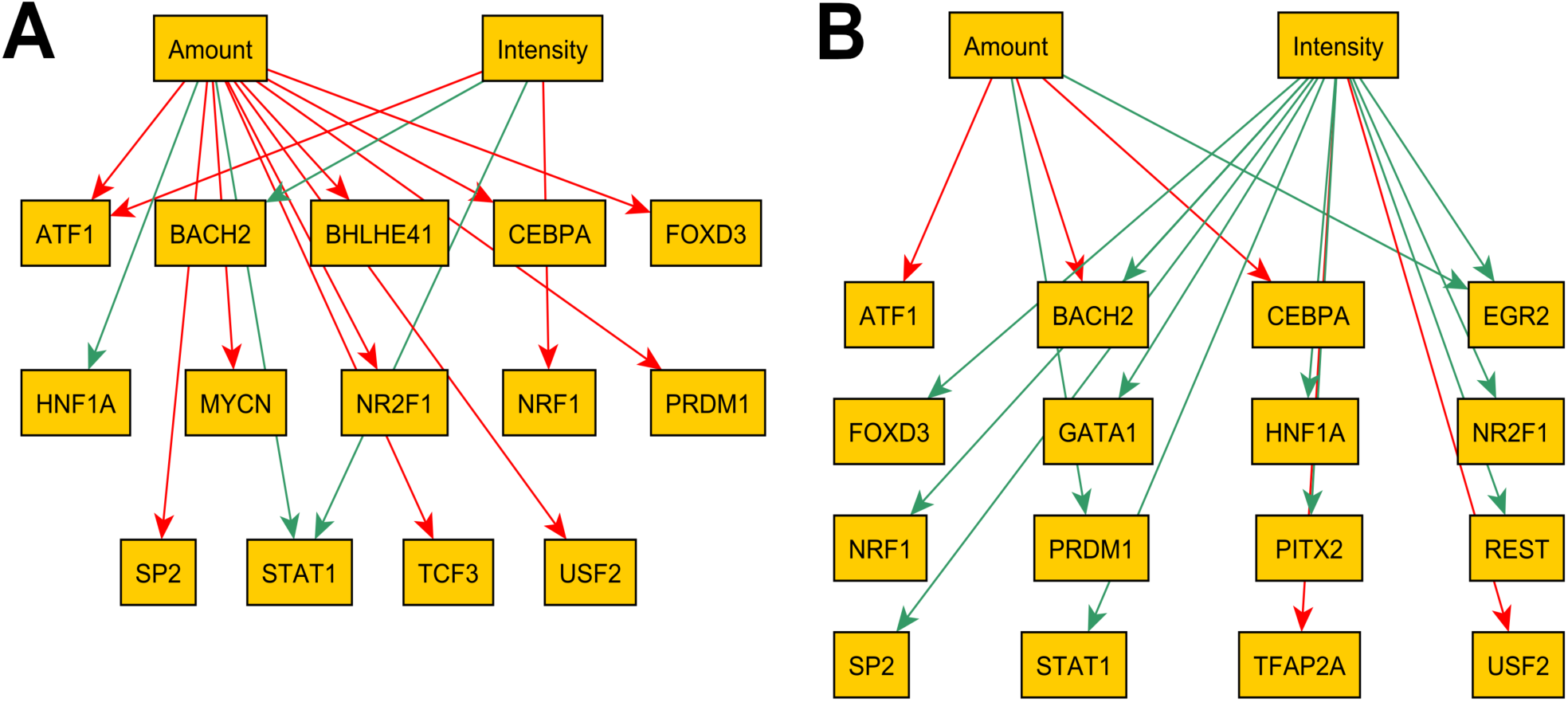
Exercise amount and intensity transcription factor gene targets; predicted direct targets of exercise amount and intensity in models for women (A) and men (B). Green edges are activating, and red edges are repressing. In women, exercise amount had substantially more targets (12) than intensity (4). In men, this pattern was reversed (5 targets for amount and 13 for intensity). The predicted effects on these targets differed as well: putative edges predominantly had a negative (inhibitory) polarity in the woman model and were concentrated on amount (10 out of 12 for amount, 2 out of 4 for intensity); in the man model putative edges predominantly had a positive (activating) polarity and were concentrated on intensity (2 out of 5 for amount, 11 out of 13 for intensity).

### Network Structure for Transcription Factor Regulation of Insulin Sensitivity

A query of the Pathway Studio database for documented regulatory interactions among the 30 identified transcription factors and three clinical measures (Si, glucose, and insulin) yielded 58 network edges, supported by 367 peer-reviewed references (minimum 2, median 3.5 per edge) (Figure 4). We then integrated regulatory actions from exercise amount and intensity into the molecular network. Both amount and intensity were potentially connected to any of the 30 transcription factors in the network, with undetermined effects (either positive or negative polarity). Amount and intensity were represented as ternary state nodes, reflecting the levels applied in the study (absent = 0, low = 1, or high = 2).

**Figure 4.**
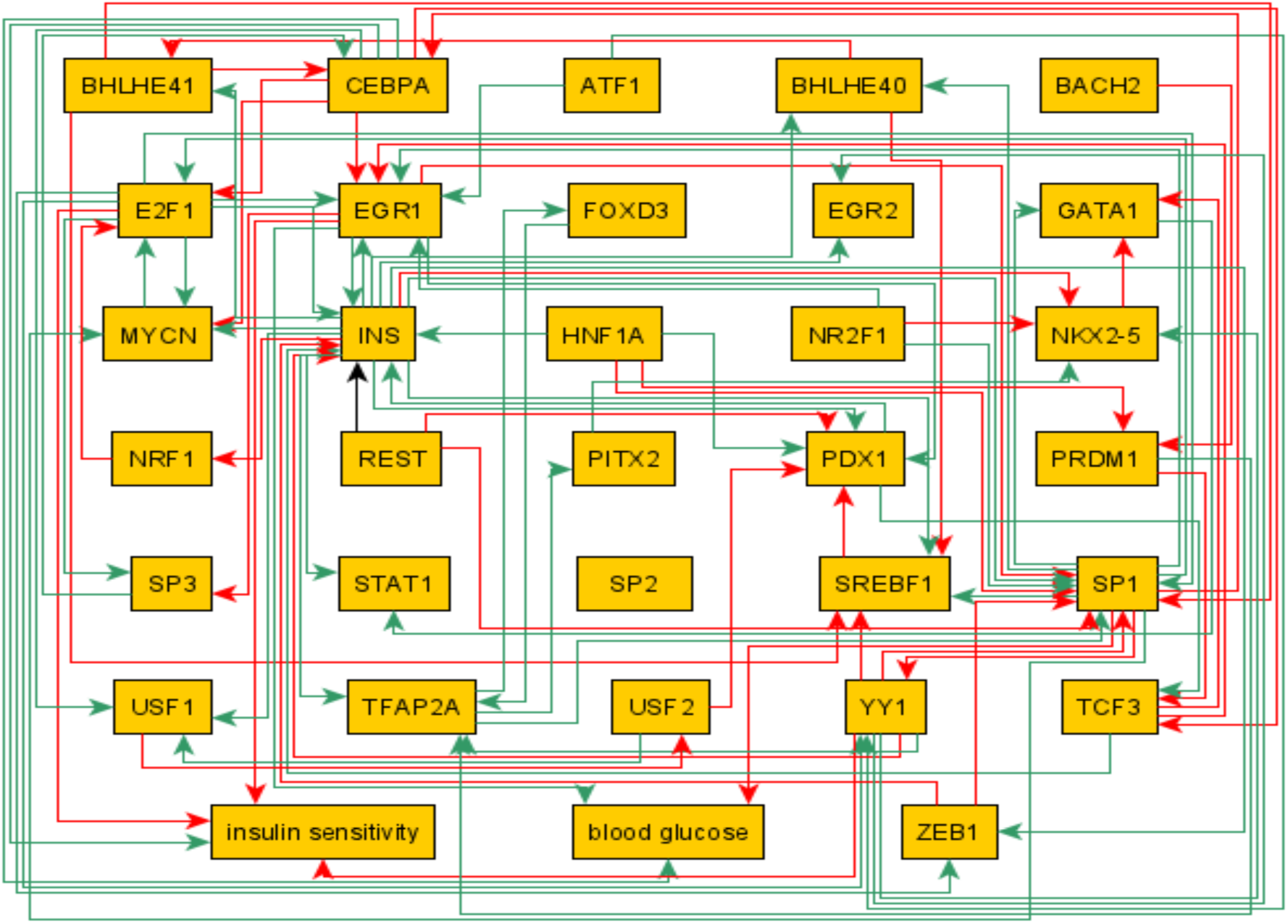
Regulatory circuit model. A transcription factor network linking 30 transcription factors through 58 documented regulatory interactions (edges) extracted from 367 full-text, peer-reviewed journal publications. Fasting insulin, fasting glucose and Si were modelled as outcomes. In this regulatory circuit diagram, green edges represent positive regulation of connecting two parameters, with the arrow pointing in the direction of the causal relationship. Red arrows are similar but indicate negative regulation. As this is a model based upon the literature, this analysis modeling regulatory circuits among transcription factor genes, it does not control for biological sex, or exercise characteristics unless so described in the literature.

### Candidate Direct Causes of Si Change (Figure 1; Step C)

We identified four Markov boundaries (sets of candidate direct causes for Si change), with each set containing four genes. RASGRP1 and the Affymetrix probe-set 236423_at (originally annotated to refseq NM_207337, now mapping to ENSG00000276900, a novel transcript antisense to AMN1) were present in all four Markov boundaries, indicating they were estimated to be direct causes of exercise-induced changes in Si. Since these two genes were identified at pre-training, this suggests a predisposition for these genes to influence training responses, possibly through genetic effects. Other variables appeared in some of the Markov boundaries and were estimated to be putative local causal factors. The cross-validation R-squared value was 0.14 ± 0.13, indicating the predictive performance of the linear regression models built with these Markov boundary variables for Si change.

Table 3 lists the variables in the four Markov boundaries and the coefficients of the linear regression models using these variables as independent predictors of Si change (post-minus pre-exercise). The coefficients indicate the expected change in Si for a one-unit change in the variable, holding the other variables in the model constant. In all models, RASGRP1 was negatively related to Si change, while probe-set 236423_at (potentially an AMN1 antisense) was positively related to Si change.

### Direct Effects of Exercise Amount and Intensity on Muscle Gene Expression (Figure 1, Step B)

We identified one Markov boundary for intensity (Table 4), indicating that the genes within this boundary are direct effects of exercise intensity. For exercise amount, we identified two Markov boundaries (Table 5). The Affymetrix probe-set 1557214_at (originally annotated as Hs.380602 and now as long intergenic noncoding RNA, LINC02382) was present in both Markov boundaries, suggesting it is likely a direct effect of exercise amount. GRIN3B and EIF3B each appeared in one of the two Markov boundaries, making them candidates for direct effects of exercise amount.

### Reconciling Literature-Based Pathway Discovery and Data-Driven Causal Discovery of Concordant Transcription Targets

In an independent analysis guided by literature-based regulatory logic models, we found that genes associated with significant changes in insulin sensitivity were regulated by 24 transcription factors targeted by exercise amount or intensity in either men or women. Among these, three transcription factors (E2F1, EGR1, and YY1) were also regulators of candidate genes directly causing changes in insulin sensitivity, but not CEBPA. Furthermore, 12 of these transcription factors were predicted to be direct targets of exercise amount and/or intensity in either the men- or women-specific regulatory models (Figure 3).

Transcription factors such as BHLHE40, E2F1, EGR1, NKX2-5, PDX1, SP1, SP3, SREBF1, USF1, YY1, and ZEB1 were not direct mediators of exercise effects on insulin sensitivity in the circuit model and were therefore excluded from the first-order (direct) effects models for both men and women. The only direct regulator of insulin sensitivity among the identified exercise transcriptional targets was CEBPA, which was predicted to be downregulated by exercise amount in both men and women. For the first-order effects models, all other regulators of insulin sensitivity were removed from the set of potential exercise targets. Table 2 lists the transcription factors commonly regulated by both sexes, even if the direction of regulation by exercise training was opposite.

### Enriched Biological Pathways of Exercise-Responsive and Insulin Sensitivity-Related Target Genes

To elucidate relationships among the genes identified using transcription factor-centric and causal modeling approaches, we explored the regulatory pathways among the transcription factors and their downstream targets revealed in the three previous analytic approaches. The expanded gene sets were used to generate the Venn diagram in Figure 5, which lists the 13 genes represented in all three expanded sets. These genes include CD3, CREBBP, EGFR, ESR1, ID2, MYC, RARA, TGFB1, TNF, TP53, and VEGF. The beta-estradiol and protein kinase C (PKC) signaling networks were identified as key connectors among transcription factors and causal modeling of Si and exercise-responsive genes (mapped in biological space in Figure 6). The strength of the phorbol ester- and estradiol-directed nodes was evident by the number of connections they had to other factors, with a notable common connection to the epidermal growth factor receptor (EGFR).

**Figure 5.**
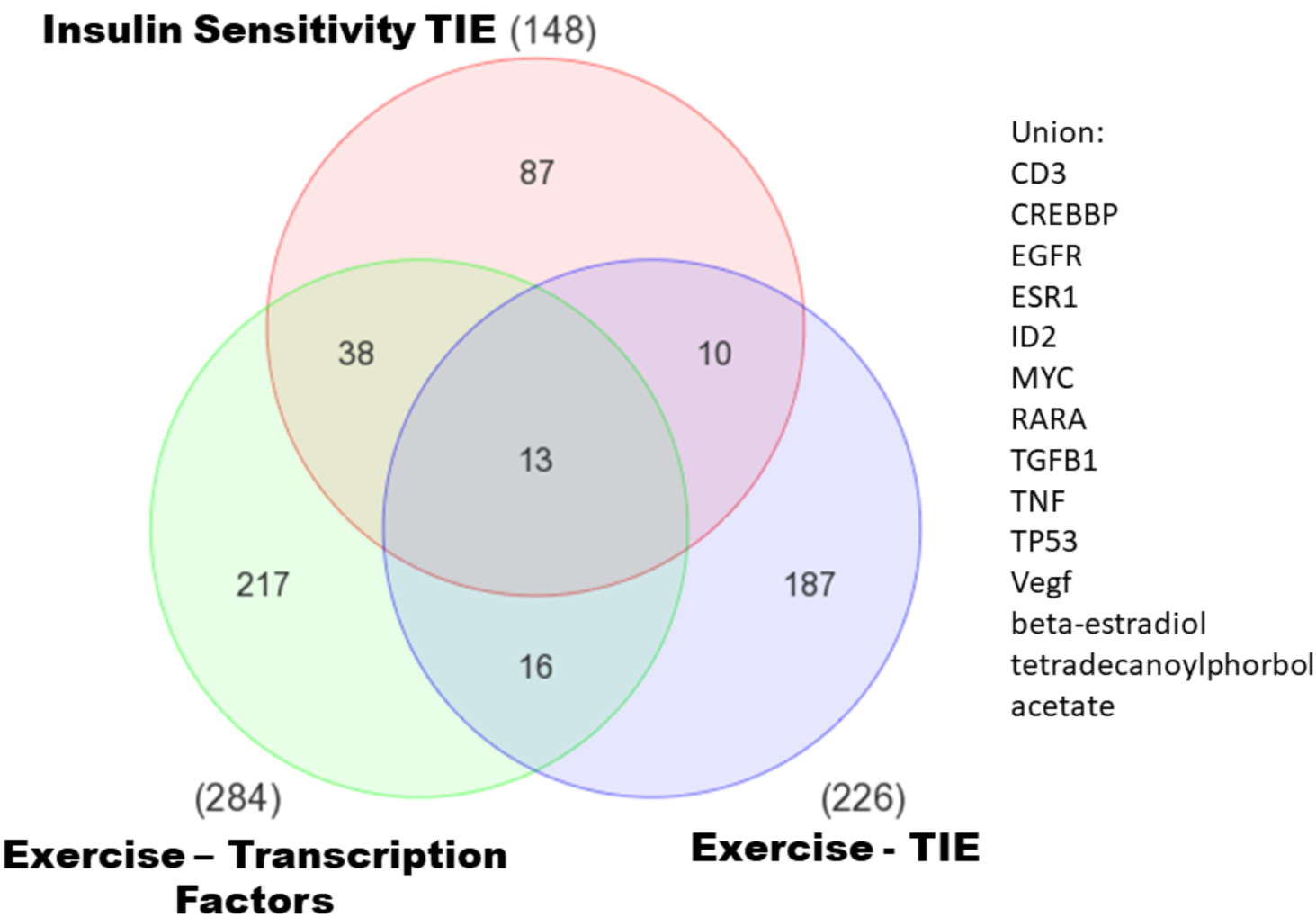
Relationships among exercise- and insulin sensitivity-related gene sets. Gene transcriptome candidates derived from Figure 1, steps A, B, and C are represented as discs in green, purple, and red respectively. Thirteen transcripts or those involved in a synthesis pathway (i.e., for beta-estradiol or tetradecanoylphorbol acetate) in common were identified. The designations are listed to the right.

**Figure 6.**
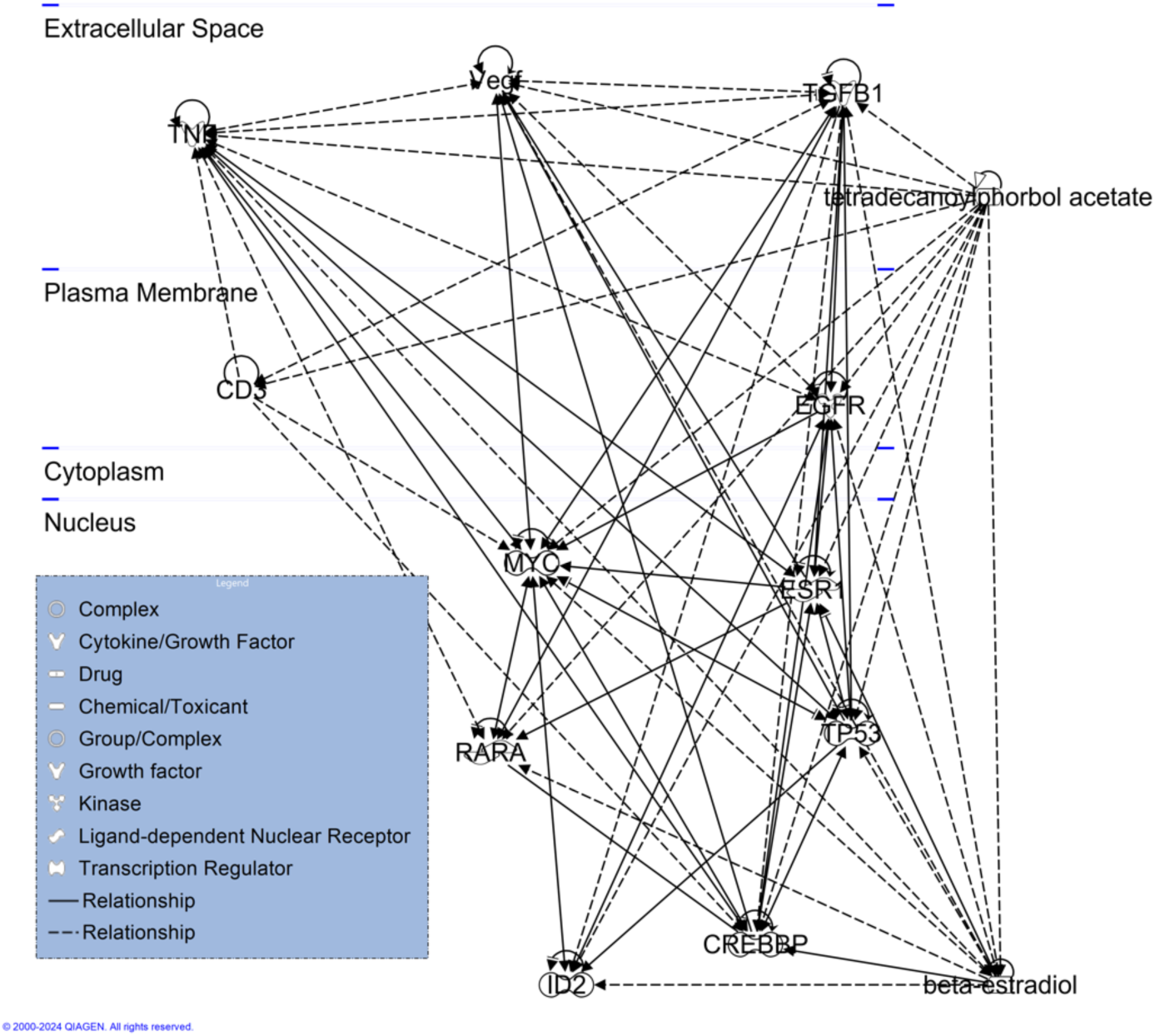
Biological relationships among exercise- and insulin sensitivity-related genes and factors identified in the analysis presented in Figure 5. In the diagram, items at the top are localized to the extracellular space, followed by the plasma membrane, cytoplasm, and nucleus (transcription factors) as one moves down the diagram and as indicated on the left. Solid arrows indicate direct molecular interactions, and dotted arrows indirect effects. Master control nodes related to phorbol ester (PKC) and estradiol signaling are seen to the far right to denote that they are not isolated to a particular subcellular compartment. Beta-estradiol, although a circulating hormone, has direct gene regulatory functions through the estrogen receptor, which are necessary for maintaining skeletal muscle fitness.^51^

## DISCUSSION

Using an innovative and newly developed analytic approach, we integrated physiological and organ-level molecular data from a prospective, randomized clinical study of exercise training with literature-driven pathway and causal modeling to create a novel and significant model. First, we identified sex-specific transcriptional pathways, responsive to exercise intensity and amount, that are causally related to exercise-induced improvements in insulin sensitivity. This model marks a significant advancement over our previous work with this dataset ^12^. Second, when considering the cumulative effects across exercise conditions, we identified two major interacting pathways: one directed by phorbol ester and PKC signaling and the other by estrogen receptor signaling. Notably, these pathways, along with two others, converged on EGFR through direct connections, highlighting EGFR as a potential pharmacologic target for enhancing Si. Additionally, other downstream factors in these pathways were identified (Figure 6).

### Sexual Dimorphism in the Transcriptional Mediators of Exercise-Induced Insulin Sensitivity (Si) Responses

While physiological differences in exercise responsiveness between men and women have been recognized for some time, the significant influence of sex on our transcription factor regulatory models was unexpected. As previously reported, both exercise amount and intensity are associated with variations in training efficacy ^13^. In this study, transcriptomic analysis of muscle biopsies collected before and after an eight-month exercise training program allowed us to identify key transcription factors linked to changes in transcriptional regulation. The predicted activity of these transcription factors at pre- and post-exercise timepoints constrained logic models designed to identify the minimal set of targets directly affected by exercise amount and intensity. Separate models for men and women yielded distinct sets of predicted transcriptional targets, suggesting that the adaptive and/or refractory effects of exercise exhibit striking sexual dimorphism.

### Interaction of Estrogen and Phorbol Ester Signaling

The role of estrogen in exercise-induced health improvements is well established. Estrogen replacement is considered crucial for aerobic exercise-induced enhancements in vascular endothelial function in postmenopausal women ^14–16^. Additionally, exercise-induced improvements in insulin sensitivity are linked to increased skeletal muscle capillarity, with sex-specific differences observed in postmenopausal women compared to age-matched men ^17^. Estrogen’s role as a major driver of exercise-induced improvements in insulin sensitivity aligns with our previous findings, where we observed an interaction between estrogen supplementation in postmenopausal women and exercise intensity on insulin sensitivity changes ^17^. In men, the differential effects of exercise intensity on insulin sensitivity were less pronounced than in women. This study confirms those findings and provides a potential mechanistic explanation.

Similarly, extensive research has established the role of phorbol ester (PKC) signaling in exercise-induced improvements in insulin sensitivity. Insulin signaling through PKC to AMP-kinase and mTOR is central to glucose transport in skeletal muscle ^18,19^. Phorbol esters affect various cellular processes, including the repression of insulin sensitivity in skeletal muscle, potentially by acting as analogs of diacylglycerol and activating PKC regulatory pathways ^19–22^. Multiple PKC isoforms are dysregulated in metabolic disease, with each subtype often having divergent roles ^23^. Recently, we reported that pharmacological PKC activation strongly induced a transcriptional signature of insulin resistance in vitro ^24^. We also observed that loss of PKC-α expression mimicked insulin resistance in vitro, while overexpression had the opposite effect. Furthermore, PKC-δ activates mTORC1, and the PKC-δ inhibitor Ruboxistaurin reverses insulin resistance in preclinical models ^25,26^. Thus, the key gene regulatory mechanisms identified in our present modeling are strongly supported by these mechanistic studies.

### Identification of Drug Target Candidates for Sex- and Intensity-Specific Effects of Exercise Training on Insulin Sensitivity

A major goal of our work is to identify molecular targets for pharmacologic agents that could mediate the health benefits of exercise. The network model presented in this study, leading to Figure 6, integrates longitudinal human clinical data, molecular data from skeletal muscle, and a novel analytic strategy—using causal modeling in a randomized prospective study—to identify potential targets. We noted that both phorbol ester and estrogen signaling converge on the epithelial growth factor receptor (EGFR) as a potential target. In fact, using a distinct modeling approach with similar objectives and different data, we also identified EGFR as a potential mediator of muscle insulin sensitivity. By integrating signatures for fasting and exercise-responsive insulin action with a drug repurposing database, we identified numerous EGFR tyrosine kinase inhibitors as pharmacologic agents that mimic insulin-related pathways in skeletal muscle in vitro ^24^. This association was later supported by extensive analyses of preclinical genetic models ^27^, and more recently, EGFR has been recognized as a therapeutic link between insulin resistance and hypertrophic cardiomyopathy ^28^. EGFR, a receptor tyrosine kinase, undergoes ligand-binding-mediated dimerization similar to insulin receptors ^29^. Amphiregulin, the endogenous EGFR ligand, represents a direct link between obesity and inflammation ^30^. Therapeutically, inhibiting EGFR enhances autophagy and insulin action ^31–33^. However, using EGFR kinase domain inhibitors to treat metabolic diseases may be challenging due to the similarity of kinase domains across the kinase proteome. Therefore, alternative protein targets upstream or downstream of EGFR that regulate its activation may represent more optimal drug targets ^24^. Furthermore, the findings from our present analysis, which account for sex-specific responses and exercise exposure characteristics (intensity and amount), provide a framework for understanding exercise dose-response relationships at a molecular level.

### Implications of this Work

The translational goal of these studies was to identify putative transcription factor targets of exercise amount and intensity in skeletal muscles, with a focus on sex-specific responses and their relationship to systemic measures of insulin, blood glucose, and insulin sensitivity. The identification of these specific targets offers promising candidates for potential intervention strategies aimed at improving insulin sensitivity in humans. However, pinpointing master regulators like estrogen and phorbol ester signaling is only the first step, as these cannot be used solely for therapies targeting Si.

For instance, while estrogen has shown beneficial effects on cardiometabolic risk factors, insulin sensitivity, and capillarity in women ^14–17^, clinical trials of estrogen supplementation in postmenopausal women did not prevent cardiovascular events or reduce mortality. In fact, some randomized trials have reported an increased risk of cerebrovascular events (stroke) with estrogen use ^34–36^. These findings highlight the potential off-target effects of systemic drug therapies intended for organ-specific outcomes, suggesting that a more practical and effective approach may involve striving for tissue-targeted effects.

Similarly, the systemic administration of PKC-signaling activators or inhibitors to improve insulin sensitivity could lead to unintended consequences. Drug targets should be more proximal, molecular, and tissue-specific, focusing directly on the site of the desired effect—in this case, Si. EGFR emerged as one such target. As identified by Timmons et al. ^24^, existing drugs can modify canonical signaling pathways involving PKC, EGFR, and mTOR, some of which could be repurposed to enhance insulin sensitivity in muscle or amplify the benefits of exercise. The regulatory networks identified in this study should be further explored to uncover other pathways amenable to pharmacologic targeting. Achieving this will require studies with larger sample sizes and broader molecular measures, such as epigenetics and proteomics, as well as investigating signaling in other organ systems like adipose tissue ^37^, liver, and pancreas ^10,38,39^, to fully assess the pleiotropic health effects of regular exercise.

### Conclusions

By integrating physiological and organ-level molecular data with literature-driven pathway and causal modeling, this study identified potential molecular targets for developing pharmacologic agents that mimic the health effects of exercise training on insulin sensitivity. Aerobic exercise-induced signaling pathways that mediate Si vary by sex and are influenced by exercise intensity and amount. The analyses provide evidence that transcriptional adaptations in skeletal muscle related to insulin sensitivity improvements are causally linked to estrogen and PKC signaling, while also accounting for sex- and exposure-related differences in exercise. By focusing on additional health benefits of regular exercise training, such as improvements in cardiorespiratory fitness, lipid metabolism, pancreatic function, and body composition, future studies using similar methods and larger sample sizes are likely to expand these findings and identify additional molecular targets. This work will critically inform the development of new therapies for the numerous health conditions that exercise effectively addresses.

## METHODS

### Study Cohort

This analysis focused on the aerobic exercise training groups from the STRRIDE I and II studies (NCT00200993 and NCT00275145) ^8,9^. This study was conducted under the oversight of the Duke University IRB; all participants agreed to participate by signing an IRB-approved consent form. Exercise training groups were categorized using a two-digit code based on the exercise program’s amount and intensity: low amount (1) of moderate intensity (1), low amount (1) of vigorous intensity (2), and high amount (2) of vigorous intensity (2). These categories were labeled as 1-1, 1-2, or 2-2, allowing us to study the effects of exercise amount while controlling for intensity, and vice versa. The exercise amount was prescribed as kilocalories expended per kilogram of body weight per week (KKW), with low amount defined as 14 KKW and high amount as 23 KKW. Exercise intensity was prescribed relative to participants’ baseline peak oxygen consumption (VtO2), assessed through a maximal cardiopulmonary exercise test. Moderate intensity was set at 40-55% of peak VtO2, and vigorous intensity at 65-80% ^8^. Participants adhered to their assigned exercise protocol for eight months, with a median adherence rate of 91.0% (IQR 79.8-99.8%).

### Participant Characteristics

The STRRIDE I and II studies recruited physically inactive adults (defined as fewer than one self-reported exercise session per week) aged 18 to 70 years, who had overweight or class I obesity (BMI 25-35 kg/m²), dyslipidemia, and metabolic syndrome, but without overt coronary artery disease or diabetes ^9^. Data for this analysis were drawn from 53 participants who had both pre- and post-training insulin sensitivity index (Si) measured using the FSIVGTT (described below), and skeletal muscle genome-wide gene expression data meeting quality control standards.

### Muscle Sampling and FSIVGTT

Muscle biopsies were taken from the vastus lateralis before the initiation of the exercise training protocol (pre) and 16 to 24 hours after the last training session (post) ^12^. Total muscle RNA was prepared as described and used for gene expression analyses ^12^. The FSIVGTT was conducted over three hours to assess blood glucose, insulin, and modeled Si at both pre- and post-training time points for all participants ^40^.

### Skeletal Muscle Genome-Wide Gene Expression

As previously described ^12^, muscle gene expression data were obtained for 39 participants using the Affymetrix HU U133 Plus 2.0 chip and for 42 participants using the Illumina HT-12 v4 Expression chip. The data were harmonized using standard NCBI gene identifiers, with 28 participants having data available on both platforms. For the Affymetrix analysis, there were five men and five women from each intervention group. A summary of participant demographics is provided in Table 1.

**Table 1.** Effects of exercise training on insulin action in study cohorts.

|  | STRIDE I & II |  | Discovery (Affymetrix) |  | Validation (Illumina) |  |
| --- | --- | --- | --- | --- | --- | --- |
|  | Pre | Post | Pre | Post | Pre | Post |
| N (m,f) | 319 (171,148) |  | 39 (21,18) |  | 42 (19,23) |  |
| Age, y | 50 ± 9 |  | 52 ± 8 |  | 51 ± 10 |  |
| Body Mass |  |  |  |  |  |  |
| Height (cm) | 179 ± 9 |  | 169 ± 10 |  | 168 ± 9.8 |  |
| Mass (kg) | 88.0 ± 13.2 | 87.0 ± 13.3 | 86.6 ± 13.4 | 85.0 ± 13.4 | 84.2 ± 10.9 | 83.8 ± 11.1 |
| BMI (kg/m<sup>2</sup>) | 30.0 ± 3.0 | 29.5 ± 3.5 | 30.4 ± 2.8 | 29.8 ± 2.7 | 29.7 ± 2.8 | 29.5 ± 2.8 |
| Insulin Action |  |  |  |  |  |  |
| S<sub>i</sub> (mU/L/min) | 3.9 ± 3.4 | 4.8 ± 4.1* | 3.8 ± 2.6 | 4.9 ± 3.2* | 4.5 ± 3.0 | 6.6 ± 5.7* |
| Change in S<sub>i</sub> (%) | 46 ± 84* |  | 44 ± 62* |  | 61 ± 88* |  |
| Fasting insulin (U/L) | 9.3 ± 5.8 | 8.2 ± 4.8* | 8.8 ± 6.1 | 7.5 ± 4.2 | 8.6 ± 6.7 | 7.5 ± 4.2 |
| Fasting glucose (mg/dL) | 94.3 ± 10.3 | 94.3 ± 9.5 | 95.0 ± 10.4 | 95.8 ± 9.7 | 92.6 ± 11.6 | 94.2 ± 9.3 |

Data are means ± SD. \* p<0.05 compared to Pre

### Overall Analytic Strategy to Identify Molecular Targets of Exercise-Induced Changes in Insulin Sensitivity

The overall analytic strategy is illustrated in Figure 1. Initially, we used a previously described approach ^41–43^ to construct a regulatory circuit model based on our entire observational gene expression dataset, which included harmonized Affymetrix and Illumina data. This model related exercise intensity and amount to changes in muscle transcription factor expression, resulting in a partially-directed causal network curated with existing knowledge from the literature using the Elsevier Pathway Studio database ^44^ (Figure 1, Step A). In parallel, we applied causal discovery Markov boundary induction methods ^45,46^ to estimate the skeletal muscle genes directly responsible for changes in Si, accounting for exercise intensity and amount, as well as the direct effects of exercise (Figure 1, Step B). The gene list generated from these analyses—constrained by the transcription factors identified in the first analysis and the causal network linking skeletal muscle gene expression to Si—was used to create a gene expression network through annotated pathway analysis software (Figure 1, Step C).

Sex-Specific Transcription Factor Targets of Exercise Amount and Intensity (Figure 2). While it is known that the expression of genes regulating insulin sensitivity differs between men and women ^6^, the sex-specific effects of exercise intensity and amount have not been fully explored. To address this gap, we analyzed transcription factor gene expression by sex. Log2 difference scores (fold-change) were calculated for each gene from pre- and post-exercise timepoints. A two-way analysis of variance (ANOVA) identified probes significantly influenced (p<0.05) by the exercise protocol, the participant’s sex, or the interaction of these variables. We then performed a multidimensional scaling (cluster analysis) to visualize the pre- to post-exercise changes in transcription factors by biological sex.

The list of significantly modified genes was submitted to PASTAA (Predict Associated Transcription factors from Annotated Affinities), an online tool that identifies transcription factors regulating differentially expressed genes ^47,48^. A p-value of <0.05 was used as the threshold for association. To calculate an “activity score” for each identified transcription factor, we multiplied the expression value of each significantly expressed gene at pre- and post-intervention timepoints by the affinity score of the associated transcription factor. These activity scores were then categorized using a gamma distribution-based expectation-maximization clustering algorithm ^49^ implemented in MatLab (Mathworks, Natick, MA). Clinical measurements of blood glucose, insulin, and Si at the pre- and post-exercise timepoints were similarly categorized. The median values for each variable within a sex group at both timepoints were used to define response trajectories by biological sex. Additional details are provided in Supplement 1.

### Simulating Regulatory Pathway Dynamics

The role of biological sex as an independent variable influencing exercise responses is not well understood, so we created separate sex-specific models. The regulatory logic of each model was adjusted independently to match the data from male and female participants, aiming to minimize hypothetical connections and use the simplest regulatory logic while maintaining a close fit to the input data, with less than a 5% departure (Manhattan distance) between the reference input and predicted output trajectories. Since only two time-separated measurements were available, the number of intermediate discrete state transitions best representing network evolution over the eight-month period was not predetermined (see Supplementary Methods).

### Data-Driven Identification of Candidate Genes Mediating Exercise Amount and Intensity Effects on Insulin Sensitivity (Figure 1, Step B)

To identify candidate direct causal genes responsive to exercise and those mediating exercise-induced changes in Si, we applied methods to identify multiple Markov boundaries ^50^ with exercise-induced changes in gene expression and Si as the target variables (see Supplementary Methods). Assuming causal sufficiency, variables present in all identified Markov boundaries were deemed true direct causal factors (true direct causes of Si or true direct effects of exercise on muscle gene expression). Variables identified in some but not all Markov boundaries may or may not have been causal, but the complete set included all true direct causal factors among the measured variables. The predictive performance of the linear regression models built with these Markov boundary variables for Si change was assessed via cross-validation.

### Pathway Analysis (Figure 1, Step C)

Genes identified through transcription factor analysis (exercise-responsive genes, Figure 1) and causal gene analysis (exercise-responsive mediators of Si, Figure 1) were further explored for enrichment in key biological pathways using Ingenuity Pathway Analysis software (Qiagen; Winter 2021 release). The genes in each of the three sets— transcription factors (Table 2), causal Si genes (Table 3), and causal exercise-responsive genes (Table 4)—were linked using the IPA Knowledge database. Since direct relationships among these genes were rare, we expanded each set by adding one additional linking node (a gene connected to two or more elements of the gene set) to enhance interconnectedness. The expanded gene sets are depicted in Figure 5 as a Venn diagram illustrating the overlap among subsets. The intersection of all three sets is shown in the pathway analysis (Figure 6).

**Table 2.** Summary of. **Figure 2: Regulatory relationships related to exercise amount or intensity consistent in men and women.**

| Source | Target | Polarity |
| --- | --- | --- |
| Amount | ATF1 | Inhibiting |
| Amount | CEBPA | Inhibiting |
| Intensity | BACH2 | Activating |

**Table 3.**
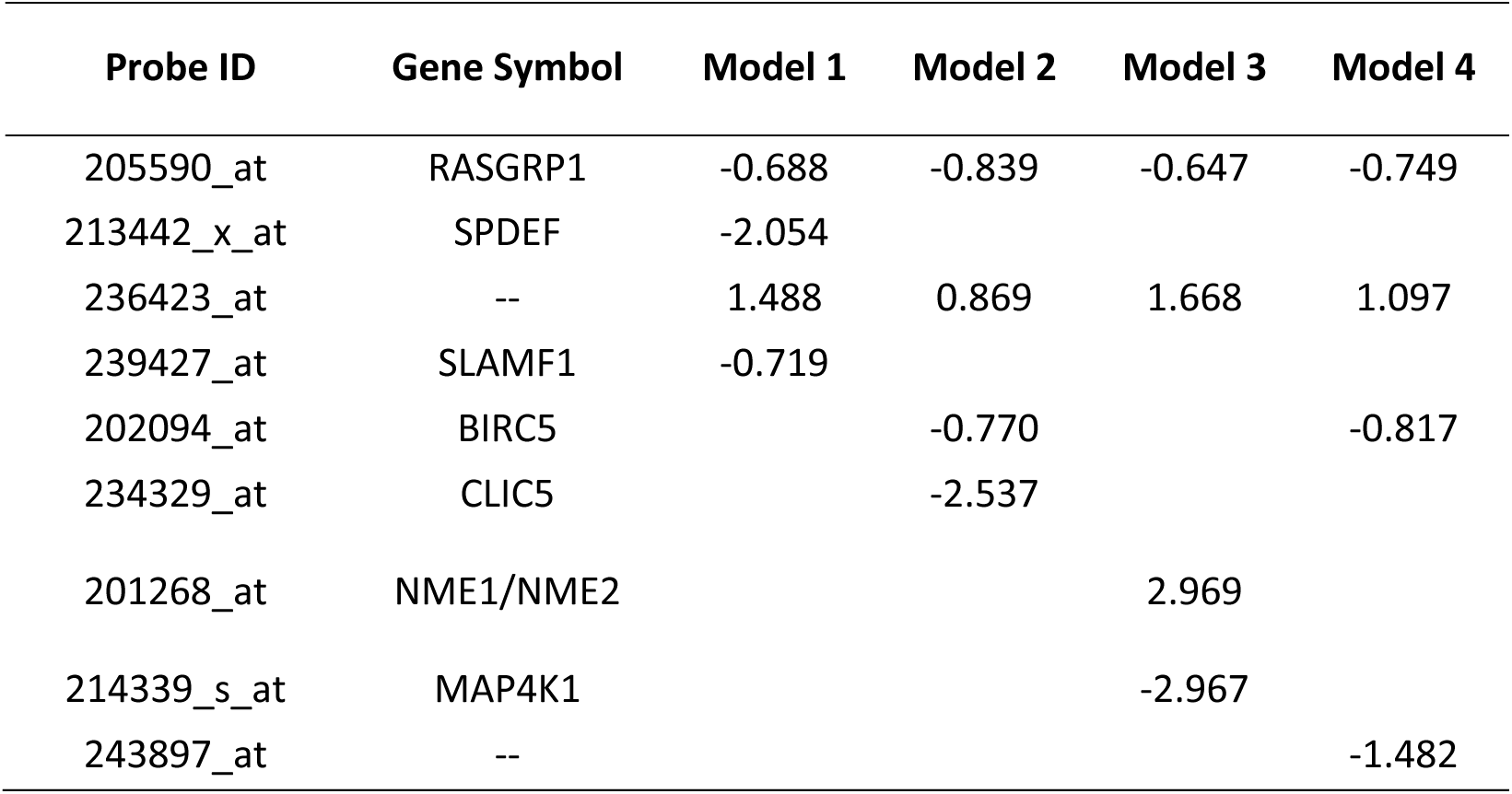
Markov boundaries for direct effects of skeletal muscle transcripts on changes in insulin sensitivity, numbers in the table shows the coefficients in the linear regression models with insulin sensitivity change as the dependent variable. By nature of the model, exercise intensity, amount and biological sex are internally controlled.

| Probe ID | Gene Symbol | Model 1 | Model 2 | Model 3 | Model 4 |
| --- | --- | --- | --- | --- | --- |
| 205590_at | RASGRP1 | -0.688 | -0.839 | -0.647 | -0.749 |
| 213442_x_at | SPDEF | -2.054 |  |  |  |
| 236423_at | -- | 1.488 | 0.869 | 1.668 | 1.097 |
| 239427_at | SLAMF1 | -0.719 |  |  |  |
| 202094_at | BIRC5 |  | -0.770 |  | -0.817 |
| 234329_at | CLIC5 |  | -2.537 |  |  |
| 201268_at | NME1/NME2 |  |  | 2.969 |  |
| 214339_s_at | MAP4K1 |  |  | -2.967 |  |
| 243897_at | -- |  |  |  | -1.482 |

**Table 4.** Expression of Markov boundary members in different exercise intensity (4a) and amount (4b) using the entire harmonized dataset.

| A. Intensity |  | Control |  | Moderate |  | Vigorous |  | F | p |
| --- | --- | --- | --- | --- | --- | --- | --- | --- | --- |
| Probe ID | Gene Symbol | n | mean (sd) | n | mean (sd) | n | mean (sd) |  |  |
| 208183_at | TACR3 | 16 | 0.72 ± 0.82 | 10 | 0.36 ± 0.55 | 23 | -0.28 ± 0.59 | 11.19 | 0.0001 |
| 1561030_at | TMC7 | 16 | -2.72 ± 4.12 | 10 | 0.81 ± 3.01 | 23 | 1.99 ± 4.87 | 5.72 | 0.006 |
| 1556422_at | --- | 16 | 0.09 ± 0.33 | 10 | 0.27 ± 0.43 | 23 | -0.29 ± 0.42 | 8.5 | 0.0007 |

| B. Amount |  | Control |  | Low |  | High |  | F | p |
| --- | --- | --- | --- | --- | --- | --- | --- | --- | --- |
| Probe ID | Gene Symbol | n | mean (sd) | n | mean (sd) | n | mean (sd) |  |  |
| 1557214_at | --- | 16 | -0.21 ± 0.67 | 23 | 0.06 ± 0.61 | 10 | 0.84 ± 0.54 | 9.13 | 0.0005 |
| 233892_at | GRIN3B | 16 | 0.22 ± 0.33 | 23 | 0.03 ± 0.40 | 10 | 0.25 ± 0.25 | 5.67 | 0.0062 |
| 237457_at | EIF3B | 16 | -2.41 ± 4.07 | 23 | -0.18 ± 3.71 | 10 | 2.44 ± 3.79 | 4.94 | 0.0114 |

## Data Availability

All data produced in the present study are available upon reasonable request to the authors

## Acknowledgements.

This study is supported by HL153497. STRRIDE I and II were supported by R01HL57354. LMR is supported by 23CDA1051777. We would like to acknowledge Eric P. Hoffman with whom both WEK and MJH worked to develop the STRRIDE gene expression work.

