## Supplemental Materials for "Sex-Specific Skeletal Muscle Gene Expression Responses to Exercise Reveal Novel Direct Mediators of Insulin Sensitivity Change"

**Appendices**

**Supplementary Tables**

**Supplement Table S1.** Participant characteristics in each aerobic exercise training group by sex and age years [summarized as median (IQR)]. Groups were low amount (1) with moderate intensity (1); low amount (1) with vigorous intensity (2); and high amount (2) with vigorous intensity (2). Participants followed the assigned exercise protocol for eight months, with a median adherence of 91.0% to their assigned intervention (IQR 79.8-99.8%). There were 5 male and 5 female participants in all intervention groups except for low amount with vigorous intensity, which had 6 male participants (Table 1).

| **Sex** | **Amount** | **Intensity** | **Count** | **Age (IQR)** |
| --- | --- | --- | --- | --- |
| F | 1 | 1 | 5 | 58.7 (53.6, 56.9) |
| F | 1 | 2 | 5 | 56.1 (46.3, 55.6) |
| F | 2 | 2 | 5 | 53.8 (49.6, 53.8) |
| M | 1 | 1 | 5 | 53.5 (51.9, 52.8) |
| M | 1 | 2 | 6 | 46.9 (46.0, 47.6) |
| M | 2 | 2 | 5 | 56.9 (51.2, 53.4) |

**Supplementary Methods**

**Transcription factor literature-based regulatory circuit analyses.** The transcription factors identified by PASTAA were used to construct a protein regulatory circuit model. Known regulatory interactions and links to blood glucose, insulin, and insulin sensitivity were recovered from the Pathway Studio database (Elsevier BV, Amsterdam), ^44^ a repository of interactions extracted from the scientific literature using the MedScan natural language processing engine.^52^ The regulatory circuit structure derived from the published literature served as a reference network model which identified novel causal connections from exercise amount and intensity effects on transcription factors were interpreted. Sets of parameter values defining logical rules, supporting regulatory dynamics, and allowing for the recovery of experimental observations, were identified using a constraint-based model-checking approach developed by our group.^53,54^ Specifically, logic parameter values were constrained to support the discretized activation scores for the included transcription factors and clinical parameters. Solutions were directed to minimize: 1) departure from the literature-derived reference data as expressed by Manhattan distance; and 2) the number of hypothetical edges included in the model and the complexity of the regulatory logic. Logic parameter identification and associated model editing were conducted incrementally across several sequential generations. Times-change was calculated by subtracting the log2-transformed pre-exercise measurements from the post-exercise measurements for all probes. Effects of sex and/or exercise amount/intensity on these changes was assessed using a two-way ANOVA.

At each generation of models, parameter estimation was performed until no new solutions were discovered within a 24-hour computing period. At the conclusion of each generation, the adjusted polarity Pa of the hypothetical new regulatory edges was calculated by: $Pa=\frac{Ep}{S}- \frac{En}{S}$, where Ep represents the number of solutions where the edge had positive polarity, En represents the number of solutions where the edge had negative polarity, and S represents the total number of solutions. Edges with a majority occurrence, |Pa|≥0.5, were accepted into the circuit model and applied with their assigned polarity in the next generation of the model, whereas edges with |Pa|≤0.05 were removed. This iterative process was continued until models for both female and male participants failed to determine or discard at least one additional edge.

We posited that confidently determined edges should have a high degree of consensus for both inclusion and polarity across a broad set of models with competing logical rule sets that all support the recovery of exercise response with equivalent accuracy. The putative regulatory action or polarity for each edge was determined by taking the difference between the fraction of all models where the edge was positive (activating) and the fraction of all models where the edge was negative (inhibiting), to account for both the polarity and the inclusion of each edge in the best-performing candidate models.

**Generation of the regulatory circuit model (Figure 3).** Simulating regulatory dynamics: simulations. After developing the preliminary regulatory circuit model, it was refined by simulations. Simulations enforcing a direct transition from the pre- to the post-exercise state resulted in the retention of all hypothetical edges by fully connecting amount and intensity to all potential molecular targets. To identify the most parsimonious actions of exercise intervention response trajectories with kinetics involving 1, 2, 3, 4, or 5 additional intermediate states (for total trajectory lengths of 3, 4, 5, 6, or 7, respectively) were tested. We found that trajectories containing 1 intermediate state enabled the discovery of solutions which produced the most significant reduction in posited hypothetical edges while preserving maximal adherence to the reference data. As the addition of further intermediate states did not significantly improve the parsimony of candidate solutions, we proceeded to conduct parameterization using reference trajectories with 1 intermediate transition state.

**Exercise amount and intensity transcription factor targets (Figure 4).** After generating the preliminary model, it was tested in further generations of simulations. After eight generations, both models failed to confidently fix or remove any additional edges (Figure. 4). At this stage, the female-context model retained 16 of the initial 60 hypothetical edges (12 from amount, 4 from intensity), and had removed 29, with 15 remaining undetermined (6 from amount, 9 from intensity). The male-context model retained 18 edges (5 from amount, 13 from intensity), and had removed 37, with 5 remaining undetermined (4 from amount, 1 from intensity). Both male- and female-context models concurred exactly on four regulatory actions, namely inactivation of ATF1 and CEBPA by increased exercise amount and activation of BACH2 and STAT1 at elevated exercise intensity (**Table 2**). Overall, 10 edges were unique to the female context, while 14 were unique to the male context. PRDM1 was targeted by amount and NRF1 by intensity in both male and female contexts, but with opposite polarity (negative in the female model and positive in the male).
